# Interpretation of mendelian randomization using one measure of an exposure that varies over time

**DOI:** 10.1101/2021.11.18.21266515

**Authors:** Tim T Morris, Jon Heron, Eleanor Sanderson, George Davey Smith, Kate Tilling

**Author notes:** Corresponding author: Tim T Morris, MRC Integrative Epidemiology Unit, University of Bristol, Oakfield House, Oakfield Grove, BS8 2BN, UK.

## Abstract

**Background:** Mendelian randomization (MR) is a powerful tool through which the causal effects of modifiable exposures on outcomes can be estimated from observational data. Most exposures vary throughout the life course, but MR is commonly applied to one measurement of an exposure (e.g., weight measured once between ages 40 and 60). It has been argued that MR provides biased causal effect estimates when applied to one measure of an exposure that varies over time.

**Methods:** We propose an approach that emphasises the liability that causes the entire exposure trajectory. We demonstrate this approach using simulations and an applied example.

**Results:** We show that rather than estimating the direct or total causal effect of changing the exposure value at a given time, MR estimates the causal effect of changing the liability as induced by a specific genotype that gives rise to the exposure at that time. As such, results from MR conducted at different time points are expected to differ (unless the liability of exposure is constant over time), as we demonstrate by estimating the effect of BMI measured at different ages on systolic blood pressure.

**Conclusions:** Practitioners should not interpret MR results as timepoint-specific direct or total causal effects, but as the effect of changing the liability that causes the entire exposure trajectory. Estimates of how the effects of a genetic variant on an exposure vary over time are needed to interpret timepoint-specific causal effects.

## Introduction

### Mendelian randomization

Mendelian randomization (MR) is used to estimate the causal effects of modifiable exposures (risk factors) from observational data under assumptions that may be more plausible than the unmeasured confounding and no measurement error assumptions required by conventional methods.^1^ MR is generally implemented within an instrumental variables (IV) framework that exploits the randomisation of genotypes at conception, using this random variation in alleles to instrument differences in observed exposures between individuals. Reverse and residual confounding are reduced because formation of genotype occurs prior to phenotypic development and is generally unrelated to environmental factors.^2,3^

Three assumptions are required for MR analyses to test the null hypothesis that an exposure *X* does not cause an outcome *Y* for any individuals. These are 1) relevance: that genotype is associated with the exposure of interest; 2) independence: that there is no common cause of genotype and outcome; 3) exclusion: that genotype does not affect the outcome through any path other than the exposure.^4^ In order to estimate an average treatment effect (ATE), we additionally assume throughout that the structural model relating IV, exposure(s) and outcome is linear and additive with homogeneous effect of exposure (at every time) on outcome.^5,6^

While it has long been recognised that MR estimates relate to exposures that generally act over a considerable period of time,^1,7^ MR studies have largely used a single measurement of exposure and outcome. Many exposures of interest vary over time,^8,9^ and time-varying genetic associations have been reported for a range of phenotypes.^10–16^ It is therefore unlikely that individuals will follow parallel exposure trajectories across the lifecourse by genotype or that consistent effect sizes will be estimated using MR applied to exposures measured at different ages across the lifecourse.^13,17^

### Mendelian randomization applied to one measure of an exposure that varies over time

It may not be appropriate to apply MR to exposures that vary over time.^18–20^ Labreque & Swanson demonstrated that MR estimates at different ages vary in the presence of time-varying genotype-exposure associations.^18^ They concluded that MR therefore provides a biased estimate of the lifetime causal effect of increasing the exposure by one unit at each timepoint throughout the lifecourse (a definition which implies time-invariant exposures and genotype-exposure associations, consistent with parallel exposure trajectories). Concerns have also been raised that MR with time-varying exposures may be biased if a feedback mechanism exists where genetic factors influence predisposition to an outcome, which in turn influence the exposure at a subsequent time point.^19^ For example, where instruments for coronary heart disease (CHD) relate to C-reactive protein (CRP) because the instruments for CHD relate to developing atheroma, which in turn increases CRP.

We propose an approach that uses MR to assess the effect on the outcome of the change in the entire exposure trajectory that would be induced by a change in genotype. To emphasise that a change in genotype affects all manifestations of the exposure, we introduce a liability *L*, which is caused by the genotype *G*, and in turn underlies all possible exposure measurements across the lifecourse. While the effect of liability on outcome is the estimand of interest, the liability is unobserved, so its effect is estimated via the measured exposure(s). Here, we consider the case with one genetic instrument (*G*), two measurements of a time-varying continuous exposure at separate occasions (*X*_0_ and *X*_1_), an outcome measured at one timepoint (*Y*), and an unmeasured confounder *U* (Figure 1). Thus, a change in genotype changes *L*, which changes both *X*_0_ and *X*_1_. The case where *X* is measured in continuous time is described in the Supplementary Text.

**Figure 1:**
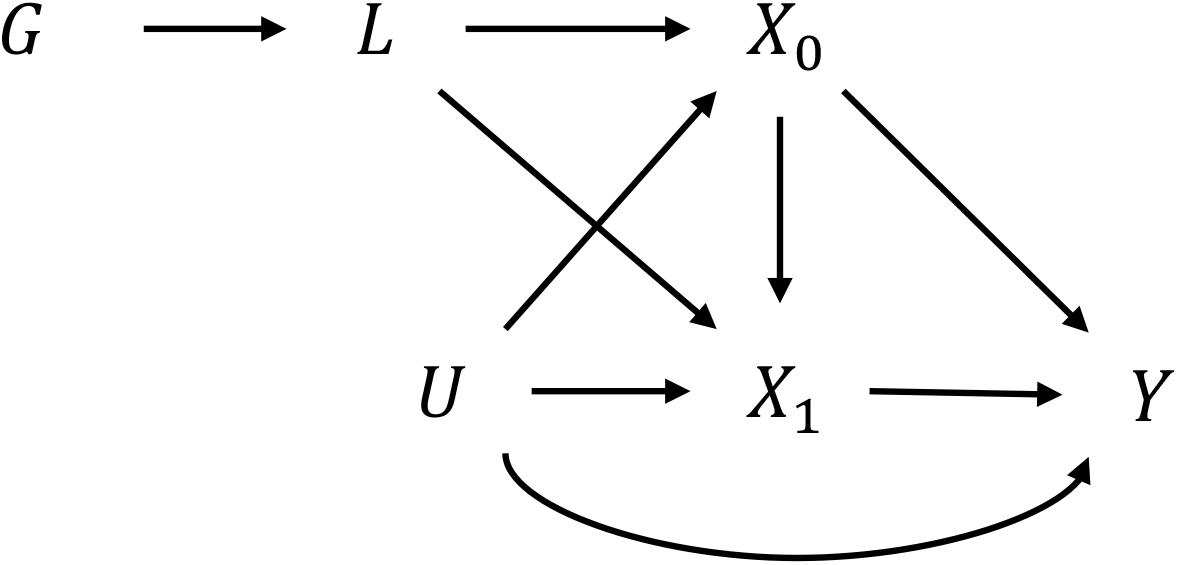
DAG showing two exposures and one outcome. ***G***, genetic instrument; ***L***, liability; ***X***_**0**_, exposure measured at time 0; ***X***_**1**_, exposure measured at time 1; ***Y***, outcome; ***U***, confounder. There is a problem of under-identification here in that the direct effects of *X*_0_ or *X*_1_ on *Y* cannot be estimated with a single liability (*L*).

Our approach overcomes two problems with interpretation of MR with time-varying exposures. First, if *G* changes, both *X*_0_ and *X*_1_ must be changed *together*; a one-unit change in *G* (e.g., an increase of one risk allele) cannot change *X*_0_ or *X*_1_ in isolation. Where time-varying genetic effects exist, the change in *G* required to raise a given exposure by one unit at time *t* (e.g., raising weight by 1kg at birth) may be different to the change required to raise the exposure by one unit at time *t* + *k* (e.g., raising weight by 1kg at age 50). Second, a one-unit change in *G* cannot have an arbitrary effect on the exposure trajectory (e.g., increase exposure by one unit at all occasions). Thus, univariable MR with one genetic instrument that acts on exposure cannot be used to recover the effect of a change in exposure at a specific time, nor of *any arbitrary* change to the trajectory of exposure. Instead, we propose that MR with a time-varying continuous exposure can be used to examine the effect of a *specific* change in the trajectory of that exposure, i.e., we are estimating the effect on the outcome of changing the exposure liability *L*.

In this paper, we clarify the causal quantities that are estimated by MR when applied to time-varying exposures with time-varying genetic effects, and how they should be interpreted. For simplicity, our example has one SNP causing *L*, though it could be proxied by multiple SNPs. The emphasis here is that the effects of *X*_0_ and *X*_1_ cannot be separated in the case where our instrument(s) act through one liability (*L*). The effects of *X*_0_ and *X*_1_ could potentially be separately estimated within a multivariable MR framework if two or more different liabilities have been identified that have different effects on *X*_0_ and *X*_1_.

## Methods

### Effects of interest

We define two estimands of interest: the total effect of a one-unit change in an exposure *X*_*k*_ (i.e. exposure *X* measured at a specific timepoint *t*_*k*_) on an outcome *Y* (*β*_*Tk*_); and the causal effect on *Y* of a change in the liability *L* that is induced by a genetic instrument *G*, such that *X*_*k*_ increases by one unit (*β*_*MRk*_). We derive algebraic expressions for these estimands in the case of two time-varying exposures and one outcome, with more general derivations given in Appendix 1.

### Total effect

We define *β*_*Tk*_ to be the total effect of *X*_*k*_ on an outcome *Y*, i.e., the change in *Y* from increasing *X*_*k*_ by one unit. This includes the direct effect of *X*_*k*_ on *Y*, and the indirect effect via the effect of *X*_*k*_ on subsequent measures of the exposure *X*_*m*_ where *m* > *k*. Note that with one liability, MR cannot be used to identify the total effect of *X*_*k*_ on *Y*.

### Liability effect of a specific genotype

We define the liability effect (*β*_*MRk*_) induced by a specific genotype as the causal effect on *Y* of changing the liability *L* such that the exposure at time *t*_*k*_ is increased by one unit. This can be thought of as the effect of moving an individual from the liability *L* giving rise to *X*= *x* at time *t*_*k*_, to a liability *L*1 that would give rise to *X*= *x* + 1 at time *t*_*k*_. This is a uniquely defined estimand for the liability associated with a specific genotype. The liability effect of *X* on *Y* will be the same for all SNPs associated with the same liability *L*, but may be different for SNPs that cause a different liability and thus a different trajectory of *X*.

We now derive expressions for the total and liability causal effect in the situation with an outcome *Y* that is caused by a genetically influenced exposure *X* measured at two timepoints (*X*_0_ and *X*_1_) (Figure 2). The genetic instrument *G* can have a non-linear relationship with the underlying liability *L*, but we assume linearity and additivity from *L* to the exposure measurements *X*_*k*_. The effect of *L* on exposure measures is allowed to change with age, thus the shape of the trajectory of *X* with age can be non-linear.

**Figure 2:**
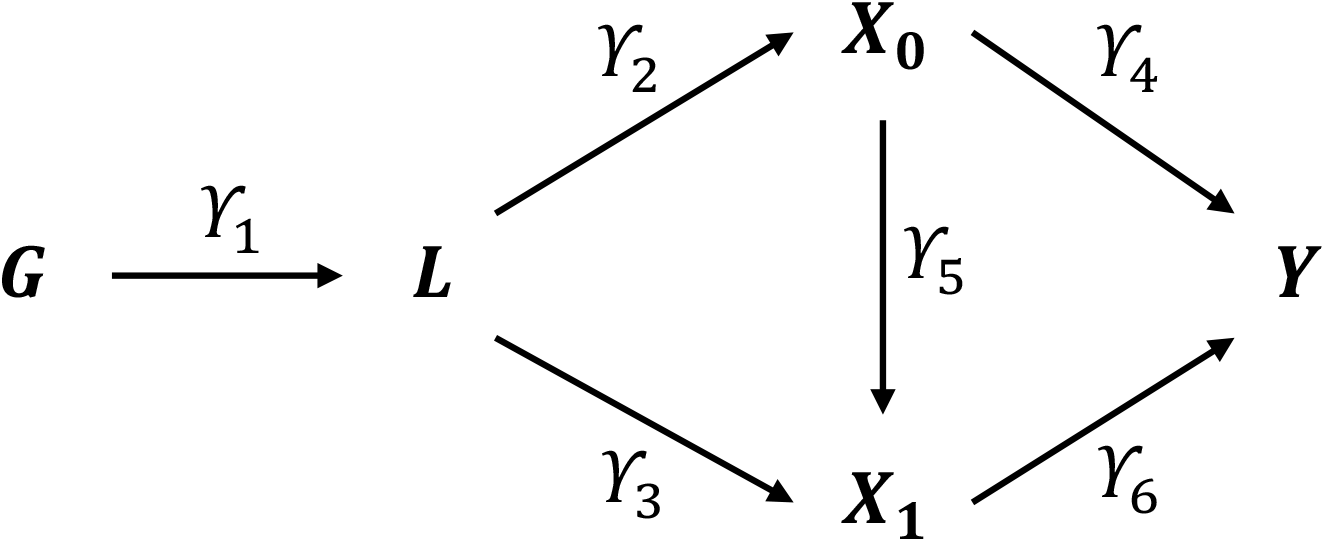
DAG showing the exposure liability in the context of two exposures and one outcome. ***G***, genetic instrument; ***L***, liability; ***X***_**0**_, exposure measured at time 0; ***X***_**1**_, exposure measured at time 1; ***Y***, outcome; ***U***, confounder.

The *total effect* of a one-unit change in *X*_0_ on 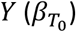 is given by:

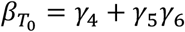

The *total effect* of a one-unit change in *X*_1_ on *Y* is given by:

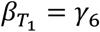

Turning to the liability effect of changes in *X*_0_ and 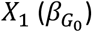, a one unit increase in *X*_0_ occurs because there is an increase in *G* from *g*_10_ to 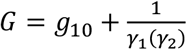

If *G* = *g*_10_ then

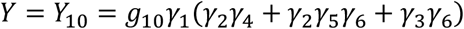

If 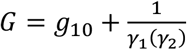 then

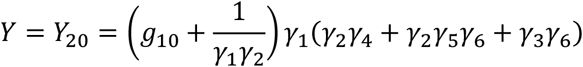

The effect on *Y* of changing the liability *L* such that it raises *X*_0_ by one unit is therefore given by:

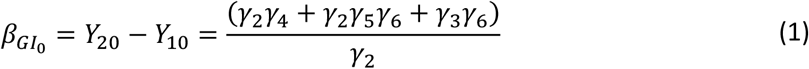

A one unit increase in *X*_1_ would occur because there is an increase in *G* from *g*_11_ to 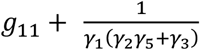

If *G* = *g*_11_ then

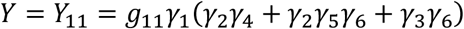

If 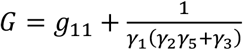 then

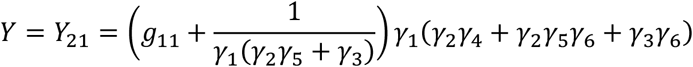

The effect on *Y* of changing *L* such that there is a one-unit increase in *X*_1_ is given by:

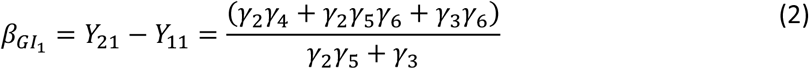

### Mendelian Randomisation

We have defined for a given SNP the liability effect (*β*_*MRk*_) as the change in *Y* induced by raising the liability *L* such that the value of the exposure *X* measured at time *t*_*k*_ is increased by one unit.

Throughout, we use the Wald IV estimator to estimate the effect of liability for an exposure that is induced by a specific genotype.

To estimate the liability effect for a given SNP of *X*_*k*_ on *Y* with MR using the Wald Ratio (*β*_*MRk*_), we need to calculate the effect of *G* on *Y*, and the effect of *G* on *X*_*k*_. In our example, we only have two measures of the exposure, so *k*=0 or 1. For examples with the exposure in continuous time, see the appendix.

The effect of *G* on *Y* is:

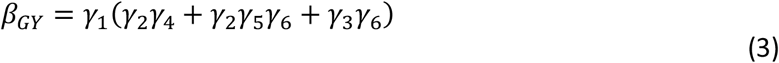

The effect of *G* on *X*_0_ is:

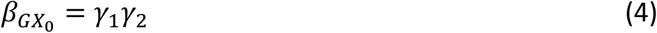

The effect of *G* on *X*_1_ is:

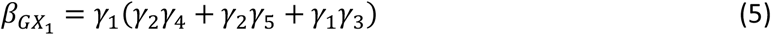

Using the Wald ratio MR estimate, the change in *Y* from changing *L* such that *X*_0_ increases by one unit (the liability effect of a specific genotype) is given by (3)/(4):

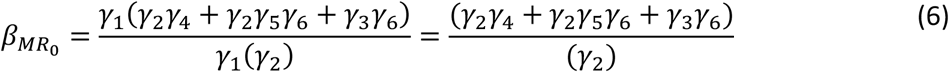

Therefore, the Wald Ratio MR estimate 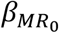 in (6) is equal to the true effect on *Y* of liability for a one-unit change in *X*_0_ in (1), and hence estimates the liability effect of *X*_0_ on *Y*.

The change in *Y* from changing *L* such that *X*_1_ increases by one unit is given by (3)/(5):

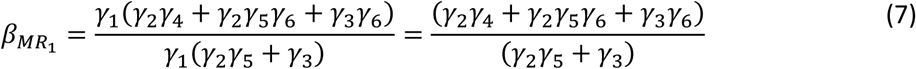

Therefore, the Wald Ratio MR estimate in (7) is equal to the true effect on *Y* of liability for a one-unit change in *X*_1_ in (2), and hence estimates the liability effect of *X*_1_ on *Y*.

MR with a single liability *L* can therefore only examine whether there is evidence for a causal effect of some measure of the exposure (at some timepoint in the period in which the liability *L* operates) on the outcome, not which part of the exposure trajectory is causal. It does not matter whether genotype-exposure associations are time-varying or time-invariant; the null hypothesis tested by MR is that the liability *L* does not cause the outcome, i.e., there is no part of the trajectory which causes the outcome. If the liability does not cause the outcome, a null effect will be correctly estimated.^19^ The Wald Ratio MR estimate of the effect of *X*_*k*_ on *Y*, the *liability effect*, is the effect of increasing *L* such that *X*_*k*_ increases by one unit. We extend this to an outcome measured at multiple timepoints in Appendix 2.

### Simulation approach

We describe our simulation approach within the ADEMP framework.^21^

#### (A)ims

The aim of the simulation was to evaluate whether the Wald Ratio is an unbiased estimator of the liability effect of *X* on *Y* for SNP *G*.

#### (D)ata-generating mechanisms

We simulated data for 10,000 hypothetical individuals (*n*_*obs*_ = 10,000), with genotypic and phenotypic data at two time points (*t*_0_, *t*_1_). *G* represents the genotype of individuals, simulated as a single variant (effect alleles = 0,1, 2) with minor allele frequency (MAF) set to 0.2 and genotype drawn from this with a binomial distribution. We simulate a time-varying exposure (*X*_*k*_) at two measurement occasions *k* (*k*=0,1), an outcome measured once (*Y*), and a time-invariant confounder (*U*) of exposure and outcome variables. Random measurement error was simulated for all variables except the genetic instrument. Base parameters were set as follows: *γ*_2_: 0.5; *γ*_3_: 0.5; *γ*_4_: 0.4; *γ*_5_: 0.3; and *γ*_6_: 0.4; (Figures 2 & 3). All associations with the unobserved confounder were set at 0.3. One-by-one these base parameters were set to zero to investigate the change in coefficient estimated. This allowed us to interrogate differential (i) strength of the genetic instrument; (ii) time-varying genetic associations; (iii) exposure effects on the outcome(s); and (iv) confounding effects. Note that the value of the unbiased estimate will not remain constant but will change depending on the base parameters. Results are presented for 1,000 replications of each simulation. All data were generated within Stata using the code available at https://github.com/timtmorris/time-varying-MR, which allows users to vary all parameters.

**Figure 3:**
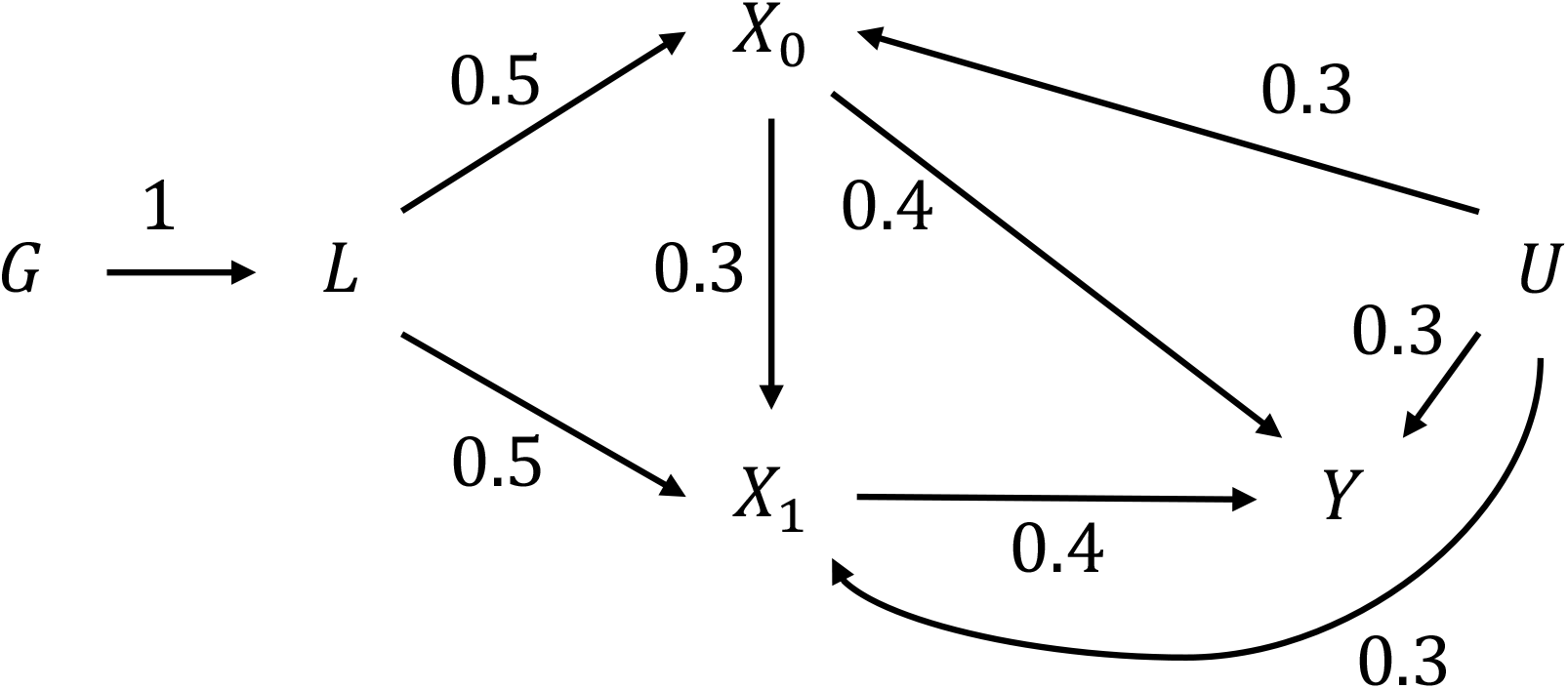
Simulated parameters. ***G***, genotype; ***L***, liability; ***X***_**0**_, exposure measured at time 0; ***X***_**1**_, exposure measured at time 1; ***Y***, outcome; ***U***, confounder.

#### (E)stimands

We estimated the causal effect of *X*_*k*_ (*k*=0,1) on *Y* by MR using the Wald ratio, and the standard error (SE) of this parameter in our simulations.

#### (M)odel

We assessed the accuracy of the Wald Ratio MR estimator.

#### (P)erformance measures

We used three performance measures: the mean of the parameter β, the mean of its SE and the deviation of β from its mathematically expected value, across the 1,000 simulations.

## Results

### Simulations

Simulations demonstrated that the Wald Ratio MR estimator correctly recovers the causal liability effect in all scenarios, even where time-varying genetic associations existed (*γ*_2_ and *γ*_3_ differ) (Table 1). Estimates of the liability effects of *X*_0_ on *Y* and *X*_1_ on *Y* differed because MR is estimating the effect of *L* on *Y* rather than the effect of *X*_*k*_ itself i.e., the change in *L* required to raise *X*_0_ by one unit is 2 (1/0.5), whereas the change required to raise *X*_1_ by one unit is 1.54 (1/0.65). Non-zero estimates were recovered for *X*_0_ on *Y* even when there was no direct path from *X*_0_ to *Y*. This non-zero coefficient arises because MR estimates the causal effect of changing the liability such that the exposure measured at time *t*_0_ is one unit higher, not the effect of a one-unit change in *X*_0_ in isolation. Non-zero effects of *X*_1_ on *Y* will be correctly estimated even if the exposure is measured after the outcome because one cannot conclude anything about temporality using MR with a single liability. MR recovered unbiased causal effects even where simulations were extended to include outcome-exposure feedback effects or reverse confounding (Appendix 2). Where the liability does not cause exposure during a specific time period (e.g. a genotype may only cause weight gain after puberty), weak instrument bias may affect estimates of the effect of the change in liability required to increase exposure by one unit during other periods (Table 1, where *γ*_2_ = 0).^22^ This bias is smaller for later measures of exposure (Table 1, where *b*_1_ = 0) because genetic effects here can operate via earlier measures.

**Table 1:**
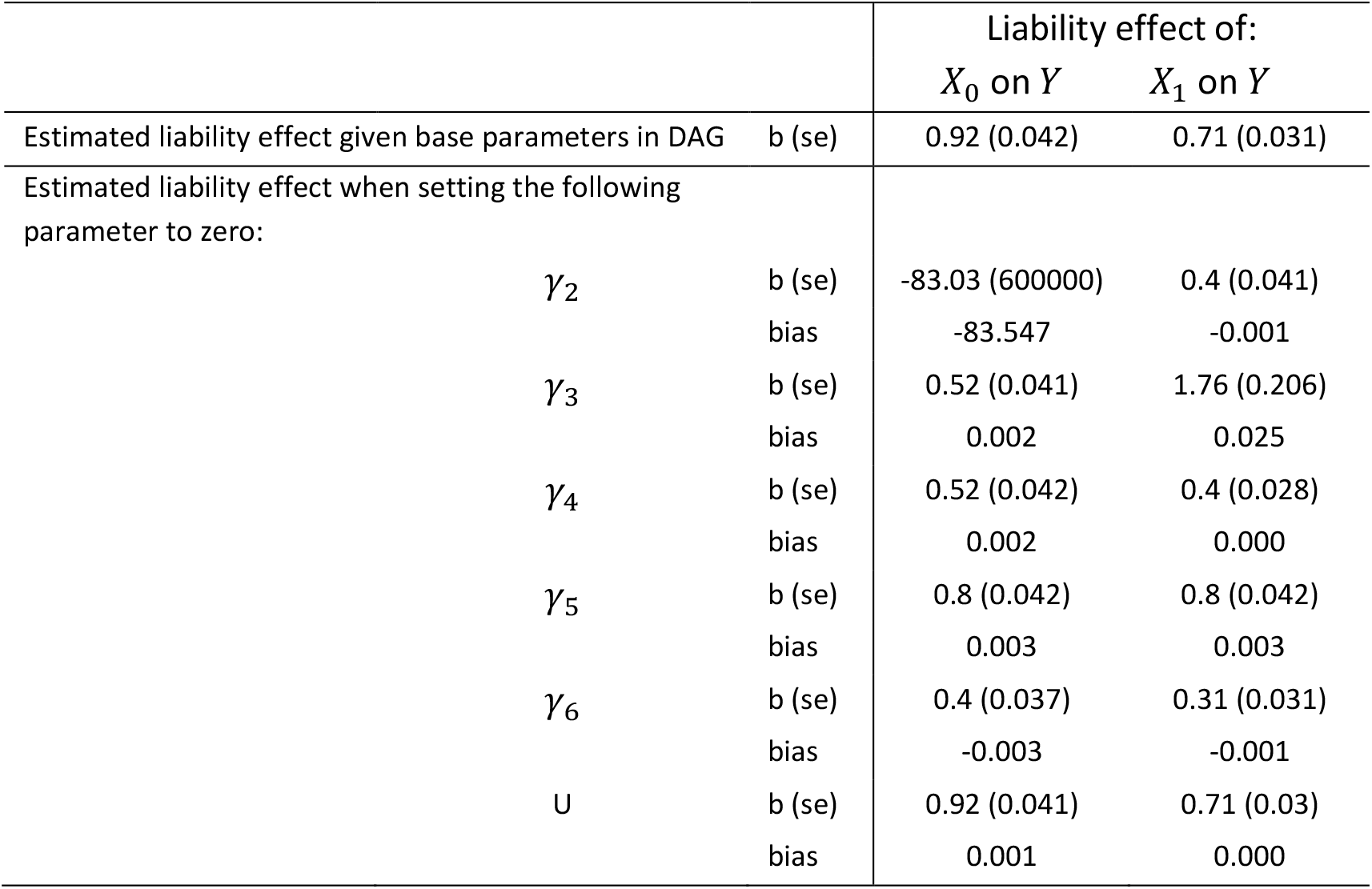
Estimates, standard errors and bias of the liability effect of a time-varying exposure on an outcome using MR. Bias presented as “0.000” where −0.001<mean bias <0.001. Note that the rows present the estimate and bias of the *target estimate* when each parameter is changed, not the estimate of the parameter itself.

Total effects estimated using linear regression were biased even where unobserved confounding from *U* was absent. This is due to confounding by the liability *L* that underlies the repeat measures of exposure (Appendix 3).

### MR of Body Mass Index measured at different ages on systolic blood pressure using FTO as the instrument

We used a two-sample MR approach to estimate the causal effect of Body Mass Index (BMI) on systolic blood pressure (SBP) using the SNP rs9939609 located in the fat mass and obesity-associated gene (FTO). Note that this single SNP approach prohibited standard 2-sample MR sensitivity analyses but provided a suitable proof of concept. We estimated FTO-BMI associations from a study using data from the 1958 National Survey of Health and Development British cohort at 11 occasions between ages 2 and 53 (n=2,479) by Hardy *et al*.^16^ We estimated FTO-hypertension associations from a study of Danish individuals in the Copenhagen General Population Study with mean age 57.6 (SD: 13.49) by Timpson *et al* (n=37,027), thus ensuring no sample overlap.^23^ All associations were consistent with the study of individuals in the Rotterdam Study (n=5,123) by Labrecque & Swanson.^18^

MR results varied greatly depending on when the exposure was measured (Table 2). This does not invalidate MR,^18^ but is expected because of the age-varying effect of genotype on exposure (Figure 4). The MR estimates should be interpreted with respect to the underlying liability; changing the liability caused by FTO such that BMI increases by one unit at age 11 would increase mid-life blood pressure by 6.08 mmHg (SE: 2.32 mmHg) (Table 2). Changing the liability caused by FTO such that BMI increases by one unit at age 53 would be to increase mid-life blood pressure by 12.78 mmHg (SE: 7.77 mmHg). Although these estimates vary, their consistency can be verified by examining Figure 4 and the effect of genotype on BMI at different ages in Table 2; the effect of a one-unit change in genotype (which would equate to a change in liability) on measured BMI is twice as large at age 11 as at age 53.

**Table 2:**
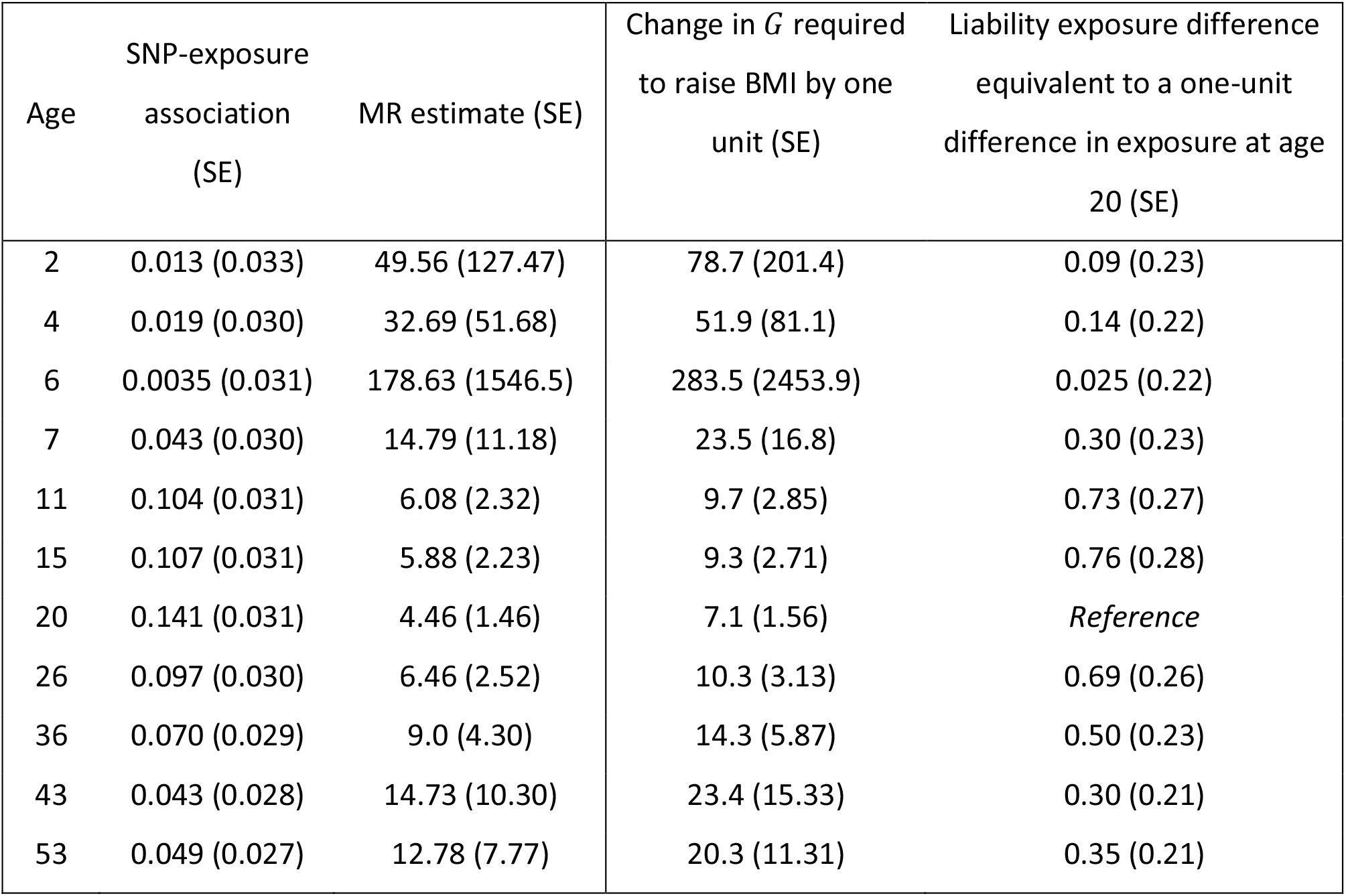
Results from MR with time-varying exposures for the causal effect of BMI z-score on SBP using FTO SNP rs9939609. SNP-exposure associations taken from *Hardy et al, 2010*;^16^ SNP-outcome association taken from *Timpson et al, 2009*.^23^ Standard errors for ratio estimates were computed using the formula in *Burgess et al (2017)*^24^ ignoring covariance between SNP-exposure effects at different ages. BMI, Body Mass Index; SBP, systolic blood pressure; SNP, single nucleotide polymorphism; MR, Mendelian randomization. The SNP-outcome association from *Timpson et al (2009)* was 0.63 (0.153).

**Figure 4:**
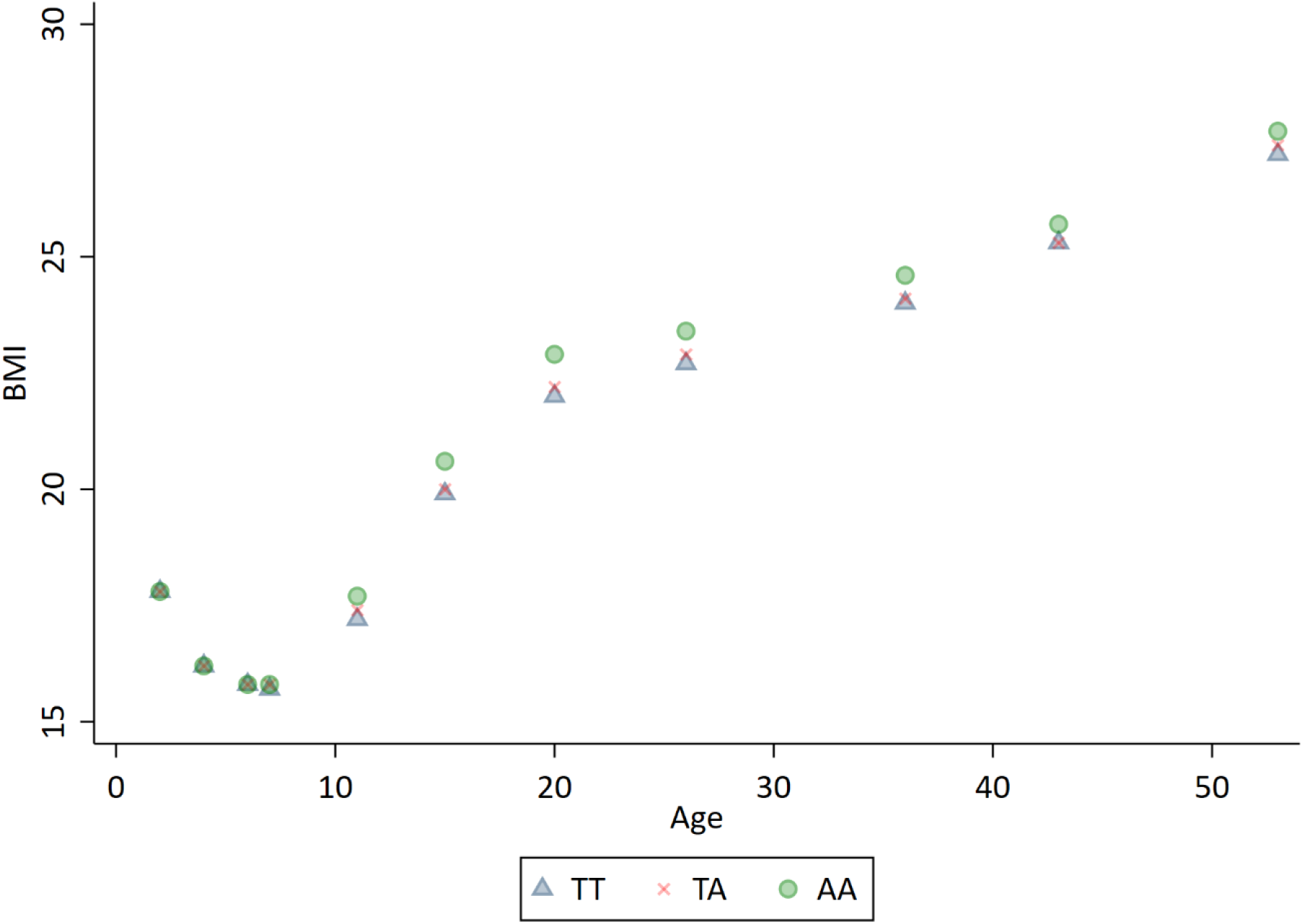
Effect of an increased risk allele on BMI at different ages from Hardy et al.^16^.

MR estimates at different exposure measurement occasions can be converted to the same liability scale provided that SNP-exposure associations at these occasions are known. This conversion is made by multiplying the Wald Ratio estimate at a reference age by the SNP-exposure association at a target age, and dividing this by the SNP-exposure association at the reference age. Taking the results in Table 2, multiplying the age 53 causal effect (12.78 mmHg per SD of BMI) by the SNP-exposure association at age 20 (0.1412) and dividing by the SNP-exposure association at age 53 (0.0493) gives us the causal effect at age 20 of 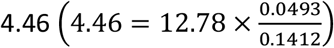. The change in *G* that would be required to raise BMI by one unit at each age can also be calculated (Table 2, column 3). This varies because a one-unit change in BMI corresponds to different genetic effects over time. The last column shows the exposure difference induced by the FTO gene at specific ages that would correspond to an FTO-induced BMI difference of one unit at age 20. This information helps to interpret the differing MR estimates and could be used to conduct a GWAS meta-analysis where the exposure was measured at different ages in different studies.

## Discussion

Here we have clarified that MR using only one measure of an exposure that varies over time gives a consistent estimate of *the causal effect of changing the liability L such that the exposure X would be one unit higher at a given time t*. That is, MR estimates the causal effect of the underlying liability rather than the causal effect of the exposure as it manifests at a given measurement occasion. Because the liability is unobserved, its effect must be estimated via measured exposures. The MR estimate of the liability effect does not require time-invariant genotype-exposure associations under the assumptions that the structural model is linear and additive, providing that the instruments are valid instruments for the underlying liability.^9,18^ There is also no assumption that the liability *L* should have the same direction of effect on exposure at all timepoints.^25^ MR with a single genetic proxy of liability cannot be used to estimate time-specific direct or total causal effects of an exposure or to draw inferences about exposure trajectories other than that caused by *L* (e.g., the effect of increasing exposure by one unit at all timepoints).^18^ MR estimates will necessarily differ by age where time-varying gene-exposure associations exist. This does not invalidate MR,^18^ but highlights that it tests the effect of the liability *L* on outcome.^26^ We have demonstrated this using two exposures for simplicity, but the result holds when the liability is extended across exposure measures in continuous time (Appendix 1).

Estimated “lifetime” causal effects may differ in size across the lifecourse, but these will be consistent with the underlying trajectory of exposure induced by the SNP. While the FTO trajectories from the study by Hardy *et al*^16^ may differ from those in larger studies,^27^ these have been used for illustrative purposes as they cover a broad range of ages. Our interpretation differs from that previously suggested^18^ in that it rests upon a liability caused by a specific genotype. Our assumption that genotype may act, through liability *L*, upon the whole lifecourse exposure trajectory^28^ rather than a single exposure measurement is supported by studies demonstrating time-varying genetic associations.^10–15^ Indeed, we argue that MR is not sensitive to age-related variation in SNP-exposure associations because these differences are a necessary component of time-varying exposures.

It may seem counter-intuitive to use the instrument to describe the causal effect to be estimated e.g., we are estimating the effect of the liability that is induced by a given SNP. The underlying point is that we can only examine the effect of a liability that has an instrument associated with it. For example, should an analyst wish to estimate the effect of increasing *X* by one unit at all timepoints, an instrument that has a constant effect on exposure over the lifecourse would be required. If the interest is in a liability that causes *X* to double every 10 years, then an instrument that has this (or a proportional) effect is required.

There are thus two consequences of our results. First, that if the aim is to estimate the effect of a specific liability, then the researcher needs to find an instrument for that liability. This is no different to any other situation where some desired exposures cannot be instrumented genetically (e.g., it is hard to imagine a valid genetic instrument for cycling to work). Second, that interpretation of an MR of an exposure that varies over time is with respect to the liability for that exposure that is induced by the given genotype. Thus, interpretation of an MR estimate of a time-varying exposure requires knowledge of the liability induced by the genotype.

Our simulations also demonstrated that MR is not biased by longitudinal exposure mediation, where earlier exposure measures cause later exposure measures. Again, it is not possible to draw inferences on the timing of causal effects because it is the effect of the liability that is being estimated, not the manifestation of the exposure at a specific time. If an outcome affects a later measurement of exposure, an investigator will not incorrectly conclude that the outcome causes the exposure, but they may incorrectly conclude that exposure at *a given age* causes an outcome. With one liability for an exposure, a cumulative effect of exposure will be indistinguishable from an effect of exposure only during sensitive or critical periods;^29^ one can only say that some part of the exposure trajectory is causal, not *which* part. The lack of ability to determine causal effects at specific timepoints complicates comparisons between MR and RCT’s, because the timing of exposure (treatment) can be modified in an RCT but randomisation of the liability underlying the exposure trajectory occurs at conception. If the mechanism of exposure is known, then the appropriate summary of exposure could be derived and used for example with cumulative exposure or functional principal component analysis to summarise trajectories of exposure.^30^

This interpretation can in principle be extended to multiple SNPs if they all act on the same underlying liability. It is highly unlikely that any two SNPs will induce exactly the same liability and thus the same exposure trajectory, but multivariable MR with multiple liabilities may allow investigators to more reliably test hypotheses about exposures during different time periods.^31,32^ For example, a recent MR study using multiple instruments with different effects on early and later life BMI could draw inferences about the different contributions of liability for BMI at ages 10 and 57.^10^ Future studies assessing heterogeneity between SNPs with repeat measures of exposure can provide valuable insight into the exposure trajectories of genetic instruments^9,18^ and better triangulate causal evidence.^33^

The key aspect when interpreting MR results from time-varying exposures is to consider the underlying liability for an exposure trajectory. Care must be taken when interpreting MR analyses using a single measure of a time-varying exposure; temporal effects cannot be inferred in the presence of a genetic instrument obtained from a single timepoint.

## Data Availability

All summary data used and code for producing the analyses are available at https://github.com/timtmorris/time-varying-MR.

## Author contributions

TTM, JH and KT conceived of the study. TTM and KT carried out all analyses. All authors contributed to interpretation of the results. TTM drafted the original manuscript. All authors contributed to editing of the manuscript. TTM will act as guarantor for the paper.

## Data availability

The summary data and all code used in this paper is available at https://github.com/timtmorris/time-varying-MR.

## Acknowledgements

We thank Tom Palmer for commenting on earlier drafts of this work.

## Appendices

## Appendix 1

### Time-varying MR extended to continuous exposure measurements

#### Lifetime effect

We wish to define a lifetime effect of *X*_*k*_ on *Y* as the change in *Y* from changing the entire trajectory of *X* such that *X*_*k*_ is raised by one unit. However, this is not a uniquely defined estimand – it does not specify how the trajectory of *X* must be changed, only that the change must be compatible with a one unit rise in *X*_*k*_. For example, the definition provided by Labreque and Swanson, of a constant one unit increase in *X* at all times, would be a lifetime effect. Other possible estimands would include the effect of changing just *X*_*k*_ by one unit, whilst keeping all other parts of the trajectory of *X* constant, or the effect of increasing *X*_*k*_ and all subsequent values of *X* by one unit. In most practical examples, none of the above estimands could be induced or observed; it is hard to imagine an RCT that could be designed to have such a specific effect on the trajectory of *X*. Thus, we must define an estimand that does have a natural and uniquely defined estimator.

We define an alternative measure of a lifetime effect as estimated by MR within the context of time-varying exposures; the effect of moving the entire exposure trajectory such that exposure at time *t* increases by one unit. This differs from the lifetime effect provided by LS, but our definition will be equivalent to theirs under the condition that individual’s exposure trajectories differ by the same quantity at every timepoint. We have an outcome *Y* which is measured only at time *T*, and an exposure *X* which is measured continuously, where *X*_*t*_ is the value of *X* measured at time *t*.

We define the liability causal effect of shifting the liability *L* (and thus the entire exposure trajectory) such that *X*_*k*_ becomes *X*_*k*_ + 1 at some time *k* as 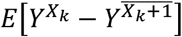. As in Figure 1, we have a genetic instrument *G* that acts on the liability *L*, which then causes *X*_*t*_. We define the instantaneous effect of *X*_*t*_ on *Y* by *γ*_*t*_; the effect of *L* on *X*_*t*_ by *β*_*Lt*_; and the effect of time on *X*_*t*_ by *β*_0*t*_:

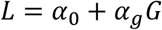

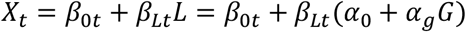

The effect of the entire trajectory on *Y* is:

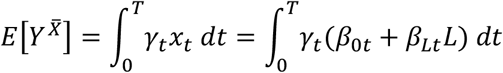

The change of the whole trajectory such that *X*_*k*_ increases by 1, means adding to *L* by 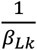

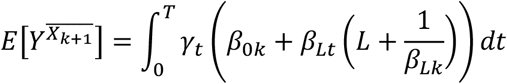

Therefore:

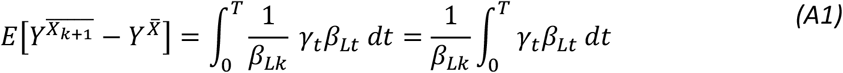

We now consider the effect on both *X* and *Y* of changes in *G*. The structural model for *X*_*t*_ when *G* = *g* is:

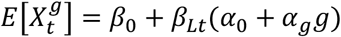

Using this in the structural model for 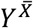:

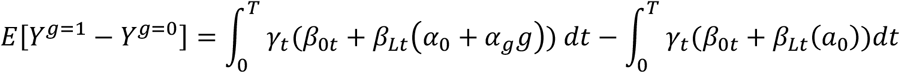

Which reduces to:

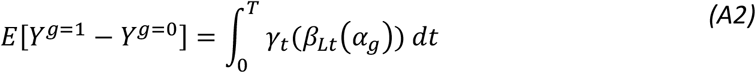

The structural model for *X* at time *k* is:

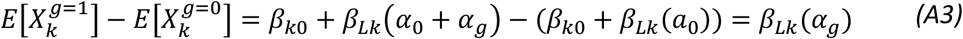

The Wald Ratio estimator is given as the change in the outcome for a given change in the instrument, divided by the change in the exposure for the same change in the instrument. Dividing the reduced form (A2) by the genetic effect measured at time *k* (A3) we obtain:

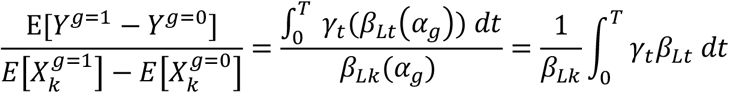

This is the same as the causal effect in equation (A1), showing that MR can be interpreted as the lifetime effect of genetically inducing an increase in exposure by one unit at time *k*.

## Appendix 2

### MR in the presence of reverse causation

#### Lifetime effect

Here we consider the case where we have a time-varying outcome (with measures *Y*_0_ and *Y*_1_) and an earlier measure of the outcome causes a later measure of the exposure (Figure A1). We show that the MR estimate is an unbiased estimate of the causal effect of a change in liability *L* such that there is a one unit change in *X* at the given time.

**Figure A1:**
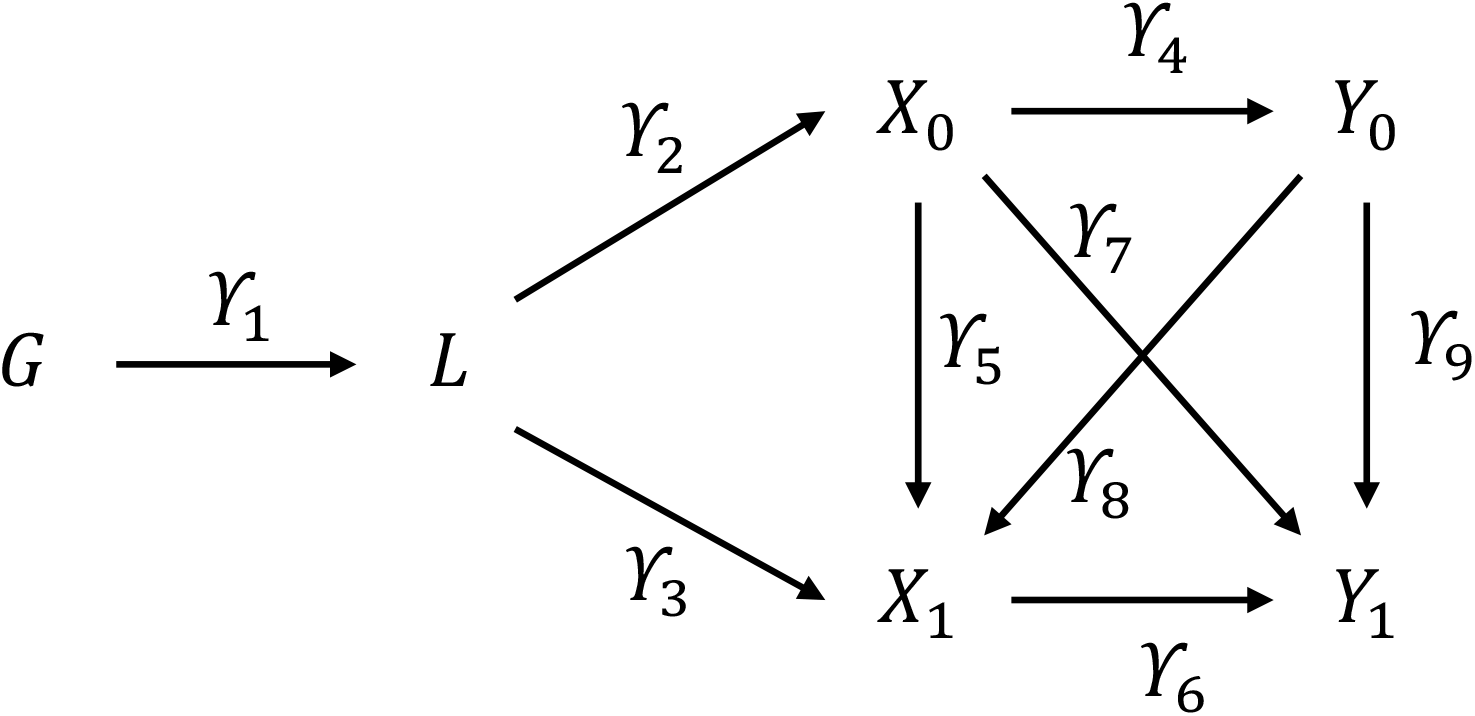
DAG showing the liability in the context of two exposures and two outcomes. ***G***, genotype; ***L***, liability; ***X***, exposure; ***Y***, outcome; ***U***, confounder. Subscripts denote timepoint.

For a given *G*, then the two outcomes are given by:

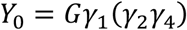

And

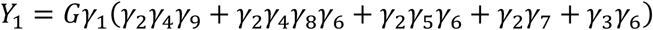

A one unit increase in *X*_0_ occurs because there is an increase from *G* to 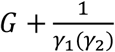

The effect on *Y*_0_ of a change in liability *L* such that there is a one unit change in *X*_0_ is therefore given by:

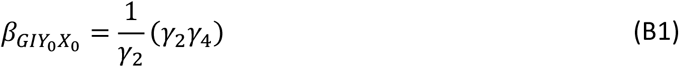

The effect on *Y*_1_ of a change in liability *L* such that there is a one unit change in *X*_0_ is therefore given by:

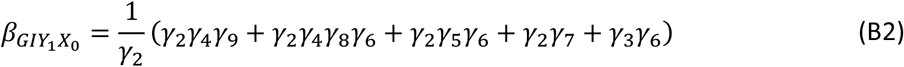

A one unit increase in *X*_1_ occurs because there is an increase from *g* to 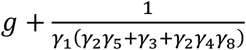

The effect on *Y*_0_ of a change in liability *L* such that there is a one unit change in *X*_1_ is therefore given by:

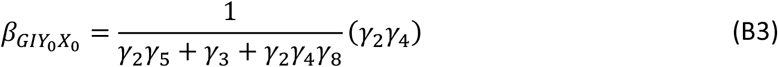

The effect on *Y*_1_ of a change in liability *L* such that there is a one unit change in *X*_0_ is therefore given by:

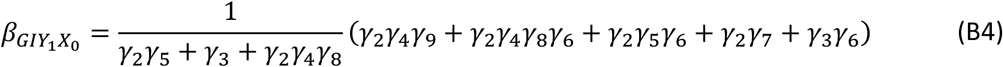

#### Mendelian Randomisation

To calculate the MR estimate of the effect of *X*_*k*_ on *Y* using the Wald Ratio, we need to calculate the effect of *G* on *Y*, and the effect of *G* on *X*_*k*_.

The effect of *G* on *Y*_0_ is:

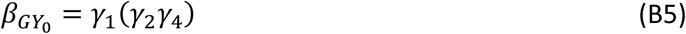

The effect of *G* on *Y*_1_ is:

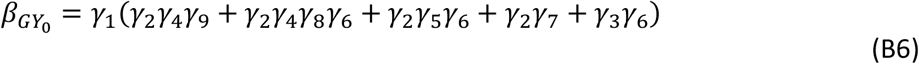

The effect of *G* on *X*_0_ is:

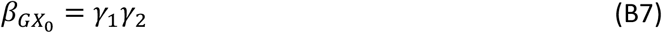

The effect of *G* on *X*_1_ is:

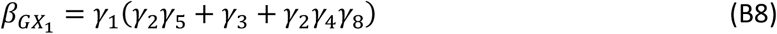

Using the Wald ratio, the MR estimate of an effect on *Y*_1_ from change in liability *L* such that *X*_0_ increases by one unit is given by (B6)/(B7):

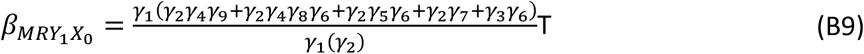

Thus, the MR estimate in (B9) is equal to the true effect on *Y*_1_ of liability for a change in liability *L* such that there is a one unit change in *X*_0_ in (B4).

Similar results follow for the MR estimates of the effect on *Y*_1_ of liability for a change in liability *L* such that there is a one unit change in *X*_1_, and on *Y*_0_ of liability for a change in liability *L* such that there is a one unit change in *X*_0_ or *X*_1_. Thus, where there is reverse causation, the MR estimates are unbiased estimates of the effect of a change in liability *L* such that *X* is one point higher at time *k*.

Simulations were repeated for two outcome measurements as shown in Figure A2. This additionally allowed us to interrogate differential effects of earlier exposures on later exposures, and reverse causation from earlier outcome measures on later exposure measures.^19^

#### Simulation approach

##### (A)ims

The aims of the simulations were to evaluate the accuracy with which MR recovers causal estimates of a time-varying exposure on a time-varying outcome.

##### (D)ata-generating mechanisms

We simulated data for 10,000 hypothetical individuals (*n*_*obs*_ = 10,000), representing a cohort sample with genotypic and phenotypic data collected at two time points (*t*_0_, *t*_1_). Let *G* represent the genotype of individuals simulated as a single variant (effect alleles = 0,1, 2) with minor allele frequency (MAF) set to 0.2 and genotype drawn from this with a binomial distribution. We simulate a time-varying exposure (*X*_*k*_) for measurement occasions *k*, an outcome measured twice (*Y*_*k*_), and a time-invariant confounder (*U*) of exposure and outcome variables. Random measurement error was simulated for all variables except the genetic instrument. Base parameters were set as follows: *γ*_2_: 0.5; *γ*_3_: 0.5; *γ*_4_: 0.4; *γ*_5_: 0.3; *γ*_6_: 0.4; *γ*_7_: 0.4; *γ*_8_: 0.2; and *γ*_9_: 0.2 (Figure A2) All confounder associations were set to 0.3. One-by-one these base parameters were set to zero to investigate the change in coefficient estimated by MR. This allowed us to interrogate differential (i) strength of the genetic instrument; (ii) time-varying genetic associations; (iii) exposure effects on the outcome(s); and (iv) confounding effects. Note that the value of the unbiased estimate will not remain constant but will change depending on the base parameters. Results are presented for 1,000 replications of each simulation. All data were generated within Stata. The programme code used to run the simulations is available at https://github.com/timtmorris/time-varying-MR and can be used to vary all parameters.

##### (E)stimands

We assessed the causal effect of *X*_*k*_ on *Y*_*k*_ and the standard error (SE) of this parameter in our simulations.

##### (M)odel

We assess the accuracy of Instrumental Variables (IV) analyses.

##### (P)erformance measures

We used three performance measures to assess the estimands in our simulations: the mean of the parameter β, the mean of the parameter SE across 1,000 replications, and the deviation of β from its expectation given the model parameters.

**Figure A2:**
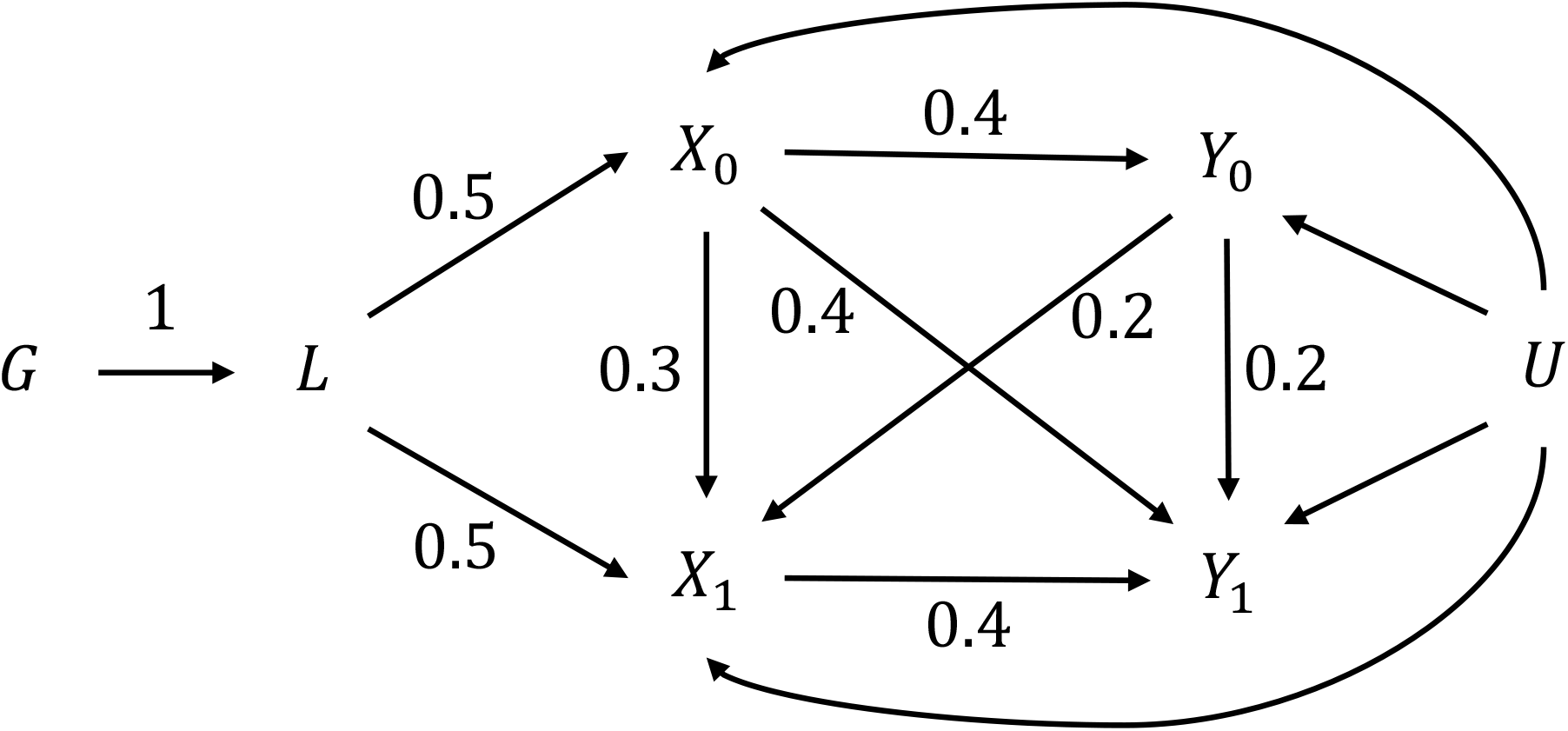
Simulated parameters. ***G***, genotype; ***L***, liability; ***X***_**0**_, exposure measured at time 0; ***X***_**1**_, exposure measured at time 1;***Y***_**0**_, exposure measured at time 0; ***Y***_**1**_, exposure measured at time 1; ***U***, confounder.

#### MR estimates of time-varying exposures in the presence of reverse causation

Simulations demonstrated that MR recovered the correct causal estimate in the presence of time-varying outcomes with outcome-exposure effects (Table A1). Where the parameter *X*_0_*X*_1_ was set to zero, a non-zero effect of 0.91 (SE: 0.04) is correctly estimated for *X*_0_*Y*_1_. Here, the effect of *X*_0_ on *Y*_1_ operates solely through *Y*_0_; both its effect on *X*_1_ and its effect *Y*_1_.

**Table A1:**
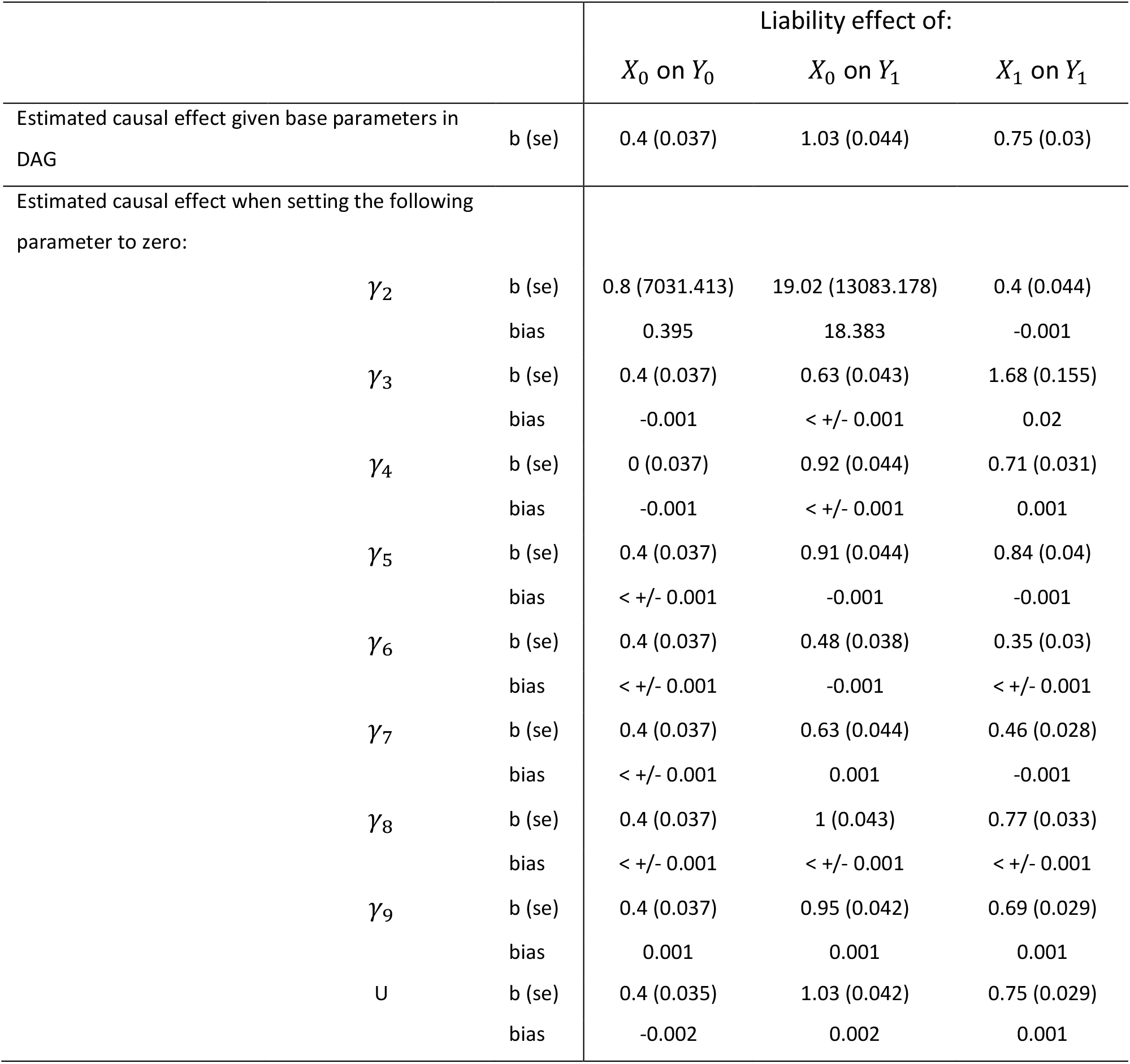
Betas, standard errors, and bias of the liability effect of a time-varying exposure on a time-varying outcome in the presence of reverse causation using MR. Bias presented as “0.000” where −0.001<mean bias <0.001. Note that the rows present the estimate and bias of the target estimate when each parameter is changed, not the estimate of the parameter itself.

## Appendix 3

### Cross-sectional total effects are confounded by genotype

Consider an outcome *Y* that is caused by two genetically influenced exposures *X*_0_ and *X*_1_ (Figure A3). Here, a linear regression model of *Y* on *X*_0_ will estimate a biased parameter for the total effect of *X*_0_ even where there is no unobserved confounding from *U*. This is because the liability underlies the repeat measures of exposure, itself acting as a source of unmeasured confounding between exposure and the outcome. This creates a condition of confounding by common intercept. A linear regression with *Y* as the dependent variable (outcome) and *X*_0_ as the independent variable (exposure) therefore estimates the total effect of *X*_0_ on *Y*, which also includes confounding by *G*. Controlling for the genetic instrument breaks this back door path of confounding in linear regression.

Linear regression estimates applied to time-varying exposures with time-varying genetic effects cannot therefore be interpreted causally, even where unobserved confounding due to traditional sources is not present. There are two circumstances in which this longitudinal confounding by liability may be avoided. First, where every other exposure measure at every other timepoint is conditioned upon, or second, where a causal effect of *X*_*k*_ exists *only* at timepoint *k*, and *X* was measured at this time. It is therefore questionable whether linear regression estimates an informative parameter in the presence of time-varying exposures.

**Figure A3:**
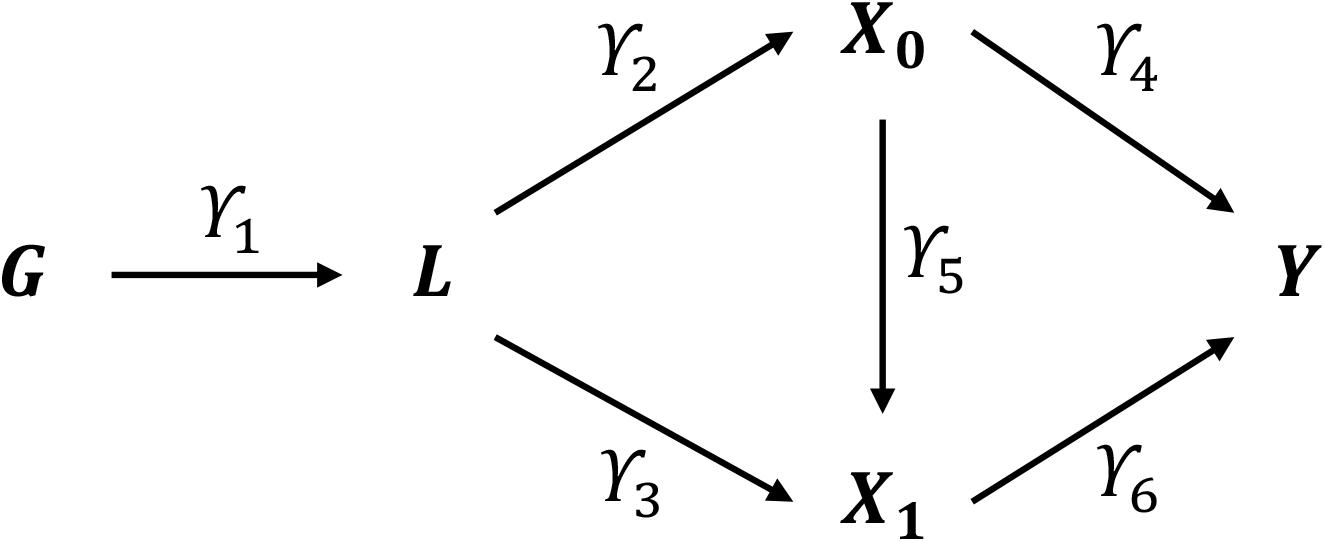
DAG showing the liability in the context of two exposures and one outcome. ***G***, genetic instrument; ***L***, liability; ***X***_**0**_, exposure measured at time 0; ***X***_**1**_, exposure measured at time 1; ***Y***, outcome; ***U***, confounder.

#### Simulation approach

##### (D)ata-generating mechanisms

We simulated data for 10,000 hypothetical individuals (*n*_*obs*_ = 10,000), representing a cohort sample with genotypic and phenotypic data collected at two time points (*t*_0_, *t*_1_). Let *G* represent the genotype of individuals simulated as a single variant (effect alleles = 0,1, 2) with minor allele frequency (MAF) set to 0.2 and genotype drawn from this with a binomial distribution. We simulate a time-varying exposure (*X*_*k*_) for measurement occasions *k*, an outcome measured once (*Y*), and a time-invariant confounder (*U*) of exposure and outcome variables. Random measurement error was simulated for all variables except the genetic instrument. Base parameters were set as follows: *γ*_2_: 0.5; *γ*_3_: 0.5; *γ*_4_: 0.4; *γ*_5_: 0.3; and *γ*_6_: 0.4 (Figure A4). All confounder associations were set to 0.3. One-by-one we changed the base parameters for *b*_1_, *c* and *u* to zero to investigate the change in coefficient estimated by linear regression. This allowed us to interrogate (i) time-varying and time-invariant genetic associations; and (ii) confounding effects. Results are presented for 1,000 replications of each simulation. All data were generated within Stata. The program code used to run the simulations is available at https://github.com/timtmorris/time-varying-MR and can be used to vary all parameters.

##### (E)stimands

We assessed the total effect of *X*_0_ on *Y* and the standard error (SE) of this parameter in our simulations.

##### (M)odel

We assess the accuracy of linear regression analyses under two approaches: (i) where the genetic instrument is omitted from the model; and (ii) where the genetic instrument is included in the model.

##### (P)erformance measures

**Figure A4:**
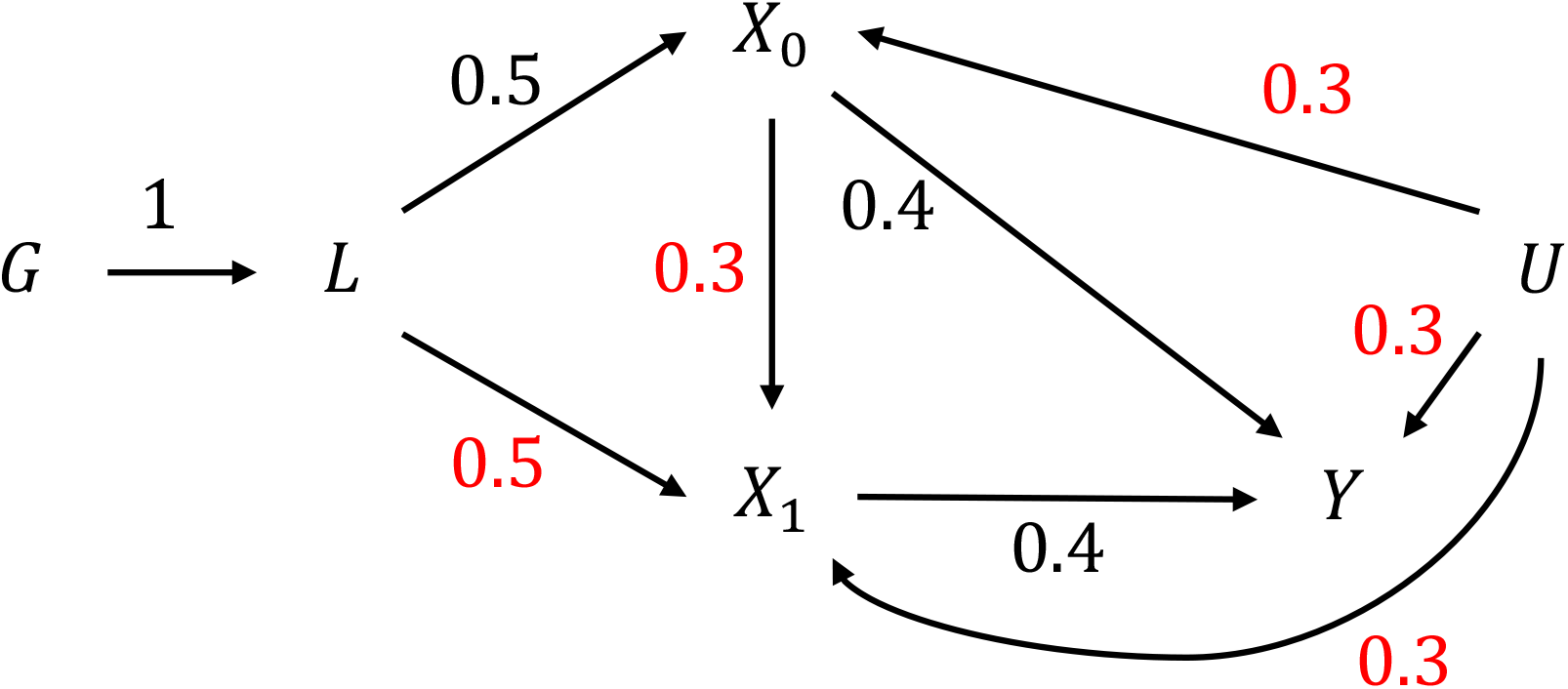
Simulated parameters. ***G***, genotype; ***L***, liability;***X***_**0**_, exposure measured at time 0; ***X***_**1**_, exposure measured at time 1; ***Y***, outcome; ***U***, confounder. Parameters in red font are those that were varied.

#### Cross-sectional total effects are confounded by genotype

Table A2 displays the results of the simulations. Linear regression failed to recover the correct total estimate of *X*_0_*Y* in the presence of unobserved confounding by *U*. Where unobserved confounding by *U* was absent, linear regression only recovered the correct total estimate of *X*_0_*Y* if the genetic instrument was included in the regression model or there was no confounding by genotype (parameter *γ*_3_ set to zero). This suggests that in the presence of time-varying genetic effects, linear regression will remain biased even where there is no unobserved confounding by traditional sources. Given the complexity of real-world exposure trajectories, this highlights the difficulty of interpreting cross-sectional estimates of a repeat measure exposure.

**Table A2:** Linear regression estimates, standard errors and bias when estimating the total effect of an exposure on an outcome (*X*_0_*Y*) using linear regression. Bias presented as “0.000” where - 0.001<mean bias <0.001.

| *Confounding*<br>*by U* | *G included in*<br>*analysis model* | *Parameter*<br>*set to zero* | *Expected*<br>*value* | *Estimate (SE)* | *Bias* |
| --- | --- | --- | --- | --- | --- |
| Absent | No | $\gamma_3$ | 0.52 | 0.520 (0.010) | 0.000 |
| Present | No | $\gamma_3$ | 0.52 | 0.628 (0.011) | 0.108 |
| Absent | Yes | $\gamma_3$ | 0.52 | 0.519 (0.011) | -0.001 |
| Present | Yes | $\gamma_3$ | 0.52 | 0.636 (0.011) | 0.116 |
| Absent | No | $\gamma_5$ | 0.4 | 0.429 (0.010) | 0.029 |
| Present | No | $\gamma_5$ | 0.4 | 0.535 (0.011) | 0.135 |
| Absent | Yes | $\gamma_5$ | 0.4 | 0.400 (0.011) | 0.000 |
| Present | Yes | $\gamma_5$ | 0.4 | 0.516 (0.011) | 0.116 |
| Absent | No | *None* | 0.52 | 0.549 (0.010) | 0.029 |
| Present | No | *None* | 0.52 | 0.655 (0.011) | 0.135 |
| Absent | Yes | *None* | 0.52 | 0.520 (0.011) | 0.000 |
| Present | Yes | *None* | 0.52 | 0.636 (0.011) | 0.116 |

